# Rapid and simultaneous identification of three mutations by the Novaplex™ SARS-CoV-2 Variants I Assay kit

**DOI:** 10.1101/2021.04.27.21255987

**Authors:** Wakaki Kami, Takeshi Kinjo, Wakako Arakaki, Hiroya Oki, Daisuke Motooka, Shota Nakamura, Jiro Fujita

**Author notes:** Correspondence to: Takeshi Kinjo, MD, PhD, 207 Uehara, Nishihara, Okinawa 903-0215, Japan.

## Abstract

**Background:** The emergence of SARS-CoV-2 variants has caused an unexpected rebound globally. The World Health Organization has listed three variants (B.1.1.7, B.1.351, and P.1) as variants of concern. To understand the epidemiology and thereby plan appropriate safety measures, differential identification of the variants is indeed critical.

**Objectives:** Although whole-genome sequencing is the gold standard for variant identification, it is time-consuming and relatively expensive. Therefore, a rapid, easy, and cost-effective platform targeting multiple regions of the genome is required. Here, we assessed the usefulness of the Novaplex™ SARS-CoV-2 Variants I Assay kit in identifying mutations in the variants.

**Study design:** We retrospectively examined 30 stored nasal swabs from COVID-19-positive patients tested between November 2020 and March 2021. RNA extracted from these swabs was subjected to the commercial kit and real-time reverse transcription-PCR was performed. To determine the genome sequences of SARS-CoV-2 in the collected samples and deduce the consensus sequences among the identified variants, genome sequencing libraries were prepared and mapped to the reference genome.

**Results:** Four of the tested samples were determined as variants. Of them, two harbored both H69/V70 deletion and N501Y substitution, whereas two harbored E484K substitution alone.

**Conclusions:** The variant with E484K substitution alone (“R.1”) has been now categorized as a variant of interest in Japan. Additionally, the kit-based assay was found to be feasible, convenient, and user-friendly in identifying the abovementioned mutations with a turnaround time of only 2 hours.

## Introduction

Although the global advent of vaccines against severe acute respiratory syndrome coronavirus 2 (SARS-CoV-2) has relieved us from the ongoing pandemic, the emergence of SARS-CoV-2 variants has raised new concerns. The World Health Organization (WHO) defines a SARS-CoV-2 variant of interest (VOI) as a phenotypically changed isolate that causes community transmission or clusters, and a variant of concern (VOC) as a VOI with increased transmissibility and virulence, or decreased effectiveness of public health and social measures or diagnostics, vaccines, and treatment [1]. As of April 13, 2021, WHO has listed three VOCs: B.1.1.7, B.1.351, and P.1, named the UK variant, South Africa variant, and Brazil/Japan variant, respectively, after the places where they were first identified [2]. Among the various mutations emerging in SARS-CoV-2, those in the spike (S) protein, specifically the H69/V70 deletion and E484K N501Y substitution mutations, are important because the S protein is involved in infectivity to host cells via angiotensin-converting enzyme 2 and the main target of neutralizing antibodies induced by vaccines [3]. All three VOCs listed above harbor the N501Y mutation; B.1.1.7 harbors an additional H69/V70 deletion; and the other two, the E484K substitution mutation. Each mutation alone and in combination determines the phenotypic features of the variants, such as transmissibility and severity of the coronavirus disease 2019 (COVID-19), and vaccine effectiveness [3,4]. Therefore, prompt differentiation of the SARS-CoV-2 variants is critical to monitor the epidemic situation in the community and to perform adequate infection control and patient care.

Currently, whole genome or complete/partial *S*-gene sequencing is the gold standard for differentiating SARS-CoV-2 variants [5]. However, genome sequencing requires professional and technical expertise, knowledge of data analysis, and high-end facilities. Additionally, it is time-consuming, laborious, and relatively expensive. Given these circumstances, a rapid, easy, and cost-effective platform targeting multiple regions of the SARS-CoV-2 genome is required [6]. Novaplex™ SARS-CoV-2 Variants I Assay is a newly developed multiplex real-time reverse transcription-PCR kit capable of identifying multiple mutations, including the abovementioned ones in a single reaction within 2 hours. Here, we report the utility of this commercial kit and discuss its implications in the differential identification of SARS-CoV-2 variants.

## Materials and Methods

We retrospectively examined the stored nasal swabs from COVID-19-positive patients tested at the Ryukyu University Hospital between November 2020 and March 2021. COVID-19 diagnosis was confirmed by real-time PCR with primer sets developed by the National Institute for Infectious Diseases of Japan [7]. RNA was extracted from the nasal swabs stored in a deep freezer using MagLEAD 12gC (Precision System Science, Chiba, Japan). The extracted RNA was then subjected to the Novaplex™ SARS-CoV-2 Variants I Assay kit (Seegene, Seoul, South Korea) according to the manufacturer’s instructions using the CFX96 Touch Real-Time PCR Detection System (Bio-Rad, Hercules, CA, USA). The test result, either plus or minus, was automatically displayed on the system’s monitor with the kit-accompanied software Seegene Viewer V1.0 (Seegene). To determine the genome sequences of SARS-CoV-2 in the collected samples, next-generation sequencing libraries were prepared using the multiplex PCR method. Briefly, after reverse transcription using SuperScript IV Reverse Transcriptase (Thermo Fisher Scientific, Waltham, MA, USA) and random primers pd(N)_6_ (Takara Bio, Shiga, Japan), whole genome amplification was conducted using ATRIC Network’s modified (V3) primer set [8]. Libraries were prepared using the Nextera XT Library Prep Kit (Illumina, San Diego, CA, USA) and 2 x 151 paired-end sequencing was performed on a MiSeq platform (Illumina, San Diego, CA, USA) with the respective MiSeq v3 Reagent kit. After mapping to the reference genome of SARS-CoV-2 (GenBank accession number: NC_045512.2), consensus sequences were obtained using CLC Genomics Workbench ver. 21.0.2 (QIAGEN, Hilden, Germany). This study was approved by the Institutional Review Board of the University of the Ryukyus (approval number: 1621).

## Results

A total of 30 COVID-19-positive samples were tested. Among them, four samples collected between February and March were determined to be variants; two (cases 1 and 2) were positive for both H69/V70 deletion and N501Y substitution mutations (UK variant), while the other two (cases 3 and 4) were positive for E484K substitution mutation alone (Table 1). These results were confirmed by whole-genome sequencing (Figure 1). In terms of usability of the Novaplex™ SARS-CoV-2 Variants I Assay kit, the procedure was simple and easy to perform, and the turnaround time was only 120 minutes (Figure 2).

**Table 1.** Result of PCR performed using the Novaplex™ SARS-CoV-222 2 Variants I Assay kit. The *RdRP* gene was included in this assay to confirm the presence of SARS-CoV-2. Endogenous internal controls of all samples were positive. (+) and (-) represent positive and negative, respectively. Abbreviations: del, deletion; Ct, cycle threshold.

|  | *RdRP* | Ct | H69/V70 del | Ct | E484K | Ct | N501Y | Ct |
| --- | --- | --- | --- | --- | --- | --- | --- | --- |
| Case 1 | ( + ) | 24.23 | ( + ) | 27.43 | ( — ) | 0.00 | ( + ) | 28.08 |
| Case 2 | ( + ) | 38.27 | ( + ) | 37.08 | ( — ) | 0.00 | ( + ) | 37.86 |
| Case 3 | ( + ) | 31.85 | ( — ) | 0.00 | ( + ) | 33.11 | ( — ) | 0.00 |
| Case 4 | ( + ) | 20.22 | ( — ) | 0.00 | ( + ) | 21.25 | ( — ) | 0.00 |

**Figure 1.**
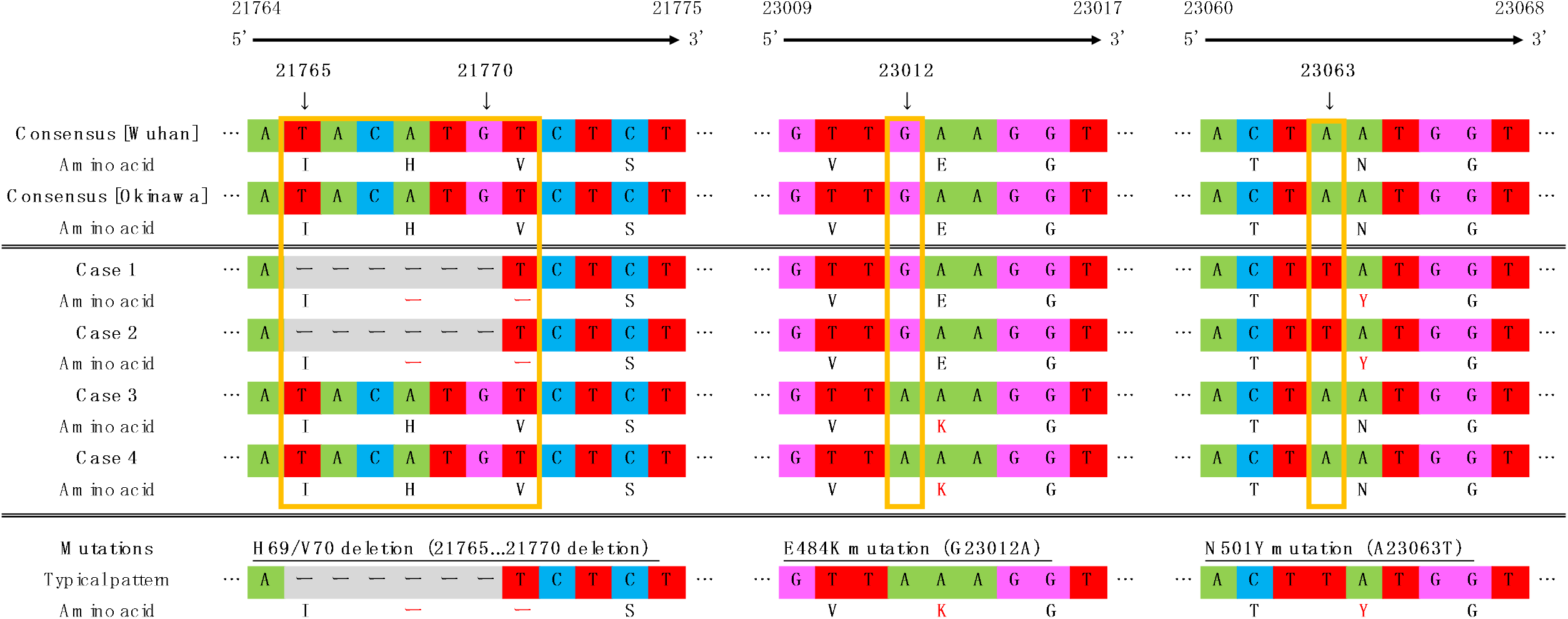
Genome sequences of the variants detected in this study.

**Figure 2.**
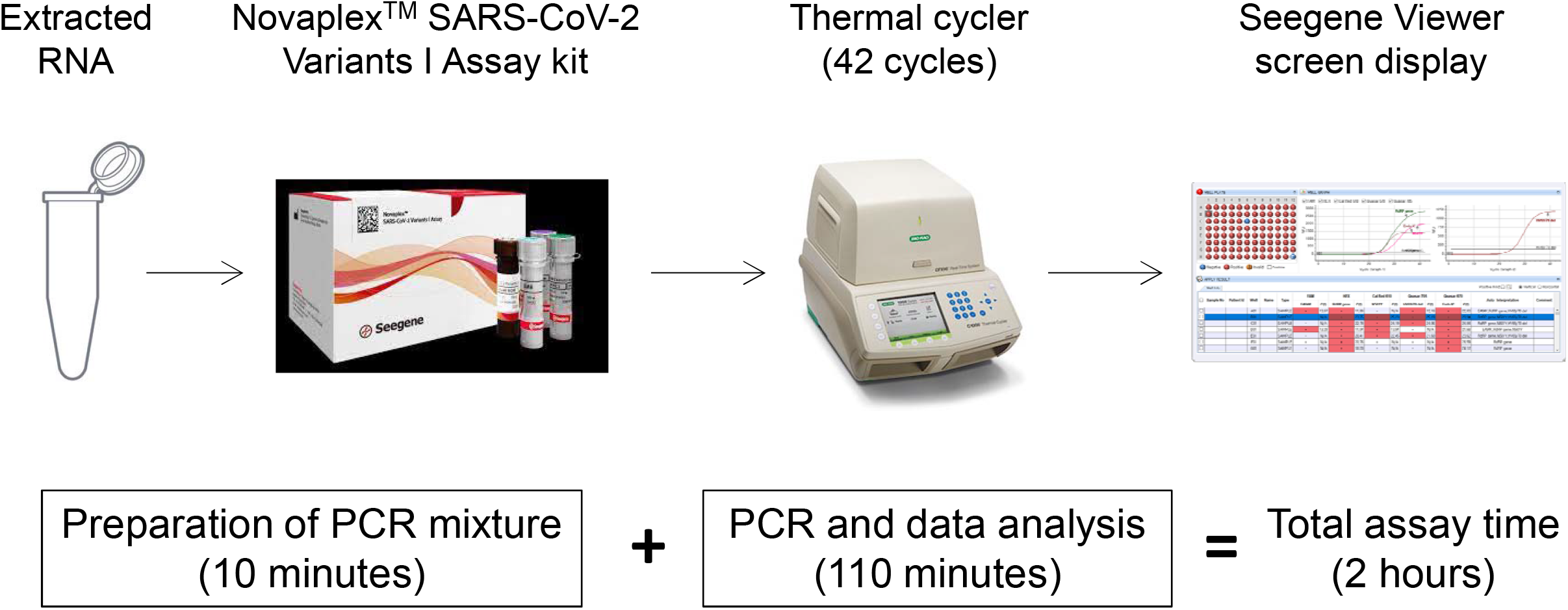
Procedure for real-time reverse transcription-PCR using the commercial Seegene kit and system.

Genome sequence data for the four cases are shown. Reference sequences (Wuhan and Okinawa) and typical mutation patterns are shown in the upper and lower stands, respectively.

PCR mixture was prepared using the extracted RNA and Novaplex™ SARS-CoV-2 Variants I Assay kit reagents; real-time reverse transcription-PCR was performed using the CFX96 Touch Real-Time PCR Detection System. The test results were automatically displayed on the monitor via the software’s interface (Seegene Viewer).

## Discussion

Many countries are now facing an unexpected rebound of the COVID-19 pandemic, mainly attributable to the emergence of several SARS-CoV-2 variants. Therefore, prompt identification of the variants is critical for controlling disease spread. Although genome sequencing is considered the gold standard for identifying SARS-CoV-2 variants, it has several limitations in terms of swiftness and convenience. In Japan, as of April 23, 2021, randomly selected COVID-19-positive samples were tested for the N501Y mutation at local public health institutes and affiliated hospitals, and the positive samples along with a few negative ones were sent to a central public health institute (National Institute of Infectious Diseases, Tokyo, Japan) for whole-genome sequencing [9]. Although dependent upon several factors, such as the local situation of the COVID-19 epidemic and geographical conditions, it usually takes a week to receive the final report of genome sequencing. This limitation can be overcome by the Novaplex™ SARS-CoV-2 Variants I Assay kit that can substantially shorten the turnaround time.

Since many variants are emerging, there is a possibility that monoplex-based testing can miss detecting some of the characteristic mutations in the variants. We detected two samples with the E484K mutation and without the N501Y mutation in this study. This variant has been named R.1 and categorized as a VOI in Japan as of April 7, 2021; however, this variant could bypass the screening stage in our country. Therefore, we need a multiplex-based test that can examine several mutations simultaneously.

In conclusion, the Novaplex™ SARS-CoV-2 Variants I Assay is a simple, rapid, and easy to perform method, and can identify three important mutations simultaneously. An additional advantage of this commercial kit is that it can be used with conventional real-time thermal cyclers. We believe that the widespread application of this kit can help in the convenient identification of the viral variants.

## Declaration of competing interest

The authors have no conflicts to declare.

## Data Availability

Data is available upon request.

## Acknowledgements

We would like to thank Editage (www.editage.com) for English language editing. We also thank Dr. Kentaro Tanaka for his technical support in performing genome analysis.

## Funding

None.

## Abbreviations

COVID-19: coronavirus disease 2019
SARS-CoV-2: severe acute respiratory syndrome coronavirus 2
VOC: variant of concern
VOI: variant of interest
WHO: World Health Organization

## Notes

### Competing Interest Statement

The authors have declared no competing interest.

### Author Declarations

This study was approved by the Institutional Review Board of the University of the Ryukyus (approval number: 1621).

